# Associations of stay-at-home order and face-masking recommendation with trends in daily new cases and deaths of laboratory-confirmed COVID-19 in the United States

**DOI:** 10.1101/2020.05.01.20088237

**Authors:** Jie Xu, Sabiha Hussain, Guanzhu Lu, Kai Zheng, Shi Wei, Wei Bao, Lanjing Zhang

**Author notes:** Correspondence: Lanjing Zhang, MD, Department of Pathology, Princeton Medical Center, 1 Plainsboro Rd., Plainsboro, NJ 08563. Conflict of Interest Disclosures: No disclosures were reported.

## Abstract

**Background and objectives:** Public health interventions were associated with reduction in coronavirus disease 2019 (COVID-19) transmission in China, but their impacts on COVID-19 epidemiology in other countries are unclear. We examined the associations of stay-at-home order (SAHO) and face-masking recommendation with epidemiology of laboratory-confirmed COVID-19 in the United States.

**Methods:** In this quasi-experimental study, we modeled the temporal trends in daily new cases and deaths of COVID-19, and COVID-19 time-varying reproduction numbers (Rt) in the United States between March 1 and April 20, 2020, and conducted simulation studies.

**Results:** The number and proportion of U.S. residents under SAHO increased between March 19 and April 7, and plateaued at 29,0829,980 and 88.6%, respectively. Trends in COVID-19 daily cases and Rt reduced after March 23 (P<0.001) and further reduced on April 3 (P<0.001), which was associated with implementation of SAHO by 10 states on March 23, and face-masking recommendation on April 3, respectively. The estimates of Rt eventually fell below/around 1.0 on April 13. Similar turning points were identified in the trends of daily deaths with a lag time. Early implementation and early-removal of SAHO would be associated with significantly reduced and increased daily new cases and deaths, respectively.

**Conclusions:** There were 2 turning points of COVID-19 daily new cases or deaths in the U.S., which appeared to associate with implementation of SAHO and the CDC’s face-masking recommendation. These findings may inform decision-making of lifting SAHO and face-masking recommendation.

---

The coronavirus disease 2019 (COVID-19) affected more than 1,125,000 people in the U.S.^1 2^. Several blind spots have been revealed and discussed. In response, many states implemented the stay-at-home order (SAHO).^3 4^ The Centers for Diseases Control and Prevention (CDC) also recommended also face-masking.^5^ Public health interventions were associated with reduction of SARS-CoV-2 transmission in China,^6-8^ but the associations of SAHO and face-masking recommendation with COVID-19 epidemiology in the U.S. are unclear. Therefore, we examined these associations using observed population data and then performed simulations for outcomes under the scenarios if early-implementation and removal of SAHO occurred.

## Methods

We extracted data about the daily new cases and deaths of COVID-19 from the COVID-19 Tracking Project, which tracked COVID-19 data since February 28, 2020^2^. Only the cases and deaths occurred from March 1 to April 20, 2020 in the 50 states and the District of Columbia were analyzed. New cases and deaths were defined as laboratory-confirmed positive cases or deaths which were reported by a state’s public health authority for the data consistency and better data-quality^2^. Each of these state authorities reported its data in different format, while most, if not all, of them followed the reporting guidelines of the Centers for Diseases Control and Prevention (CDC). It is noteworthy that on April 14, 2020, the CDC updated its definition of positive cases and included probable-positive cases.^9^ The impact of this change to the data released on April 20, 2020 would be minimal due to the short time-interval (many states have not adopted the criteria yet) and the confirmatory laboratory-test result. Therefore, the case/death numbers reported here might be smaller than those reported by others. For quality control of the released and curated data, the COVID-19 Tracking Project employed a 4-tier score system, which included 4 simple components, namely, reporting positive test results reliably, reporting negatives sometimes, reporting negatives reliably and reporting all commercial tests. Based on the sum of these scores, each state corresponded to a letter grade A, B, C and D, with A for the best quality. All states scored A or B. The study was exempt from the review by an Institutional Review Board for the use of publicly available de-identified data.

Several population-based factors were included in the multivariable piecewise log-linear regression analyses. Specifically, the timing of SAHO and populations of the states were obtained to calculate the number of subjects and the proportion of the U.S. population under SAHO on a given date.^3^ The state populations were extracted from the U.S. Census (up to July 2019).^10^ The proportions of daily positive results in all daily tests, and State-level daily new cases and deaths were obtained from the COVID-19 Tracking Project.^2^

The time-varying reproduction numbers (Rt) were defined as the mean number of secondary cases generated by a typical primary case at the time t in a population, and estimated using previously-reported serial-intervals^11-13^and the R package (Version 3.6.3).^6 14^ Three-day moving averages of the Rt were reported for their better sensitivity than 5-day moving averages. We estimated the segmental coefficients using piece-wise log-linear models and 2 presumptive turning-points. Simulation studies were performed using the prediction function. Statistical analyses were performed using Stata (version 15) and the Joinpoint program (NCI, version 4.7.0.0) with the Poisson Variance option. All *P* values were 2-sided, with a cutoff of 0.05 for significance.

## Results

On March 19, 2020, the State of California started a stay-at-home order (SAHO) which affected 3,9512,223 (12.0% of the U.S. population) U.S. residents (**Figure 1**). The number and the proportion of U.S. residents under SAHO continued to increase until April 7, and plateaued at 290,829,980 and 88.6%, respectively, afterwards.

**Figure 1.**
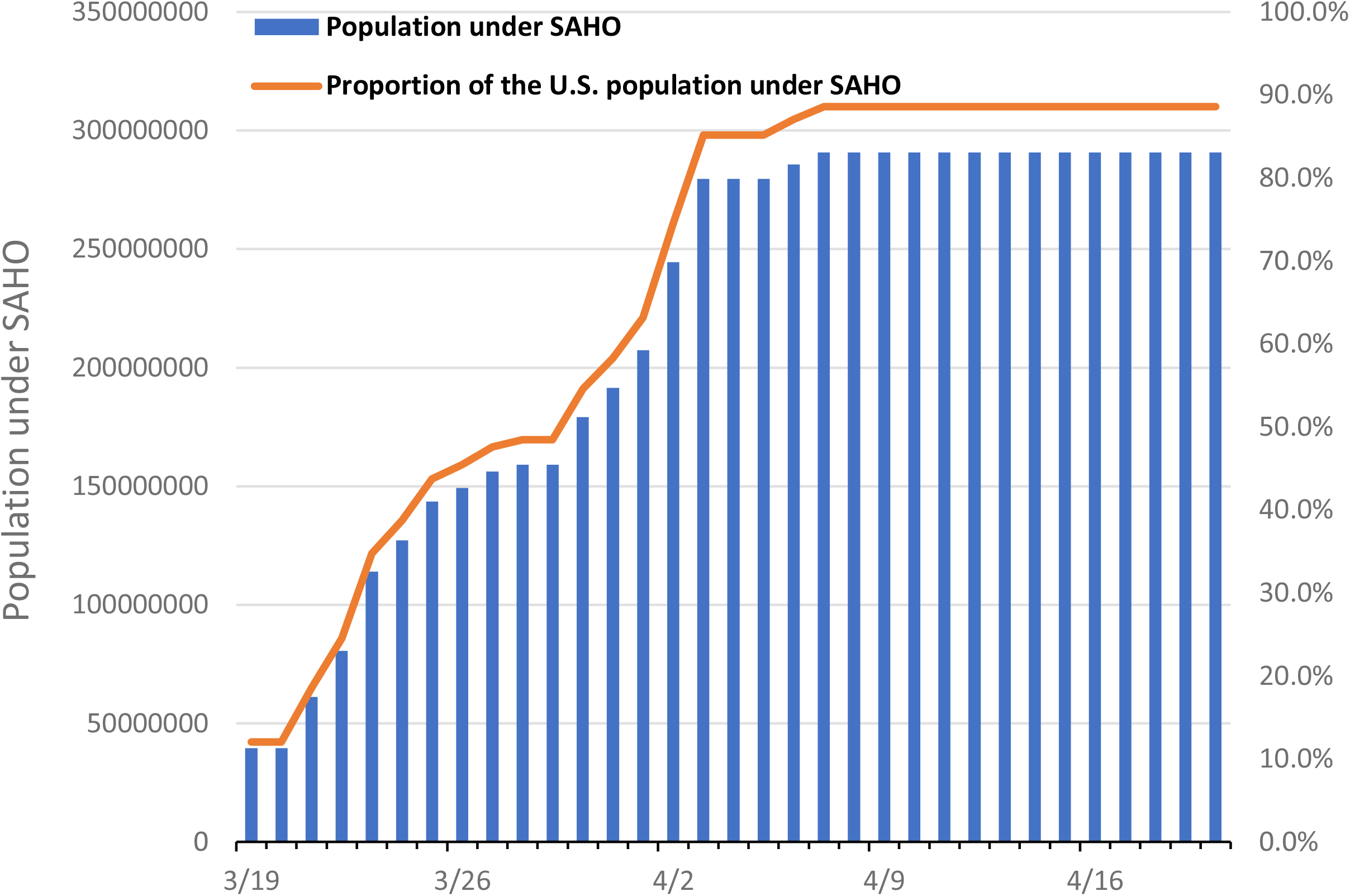
The population under a stay-at-home order owing to the COVID-19 in the United States. Since March 19, 2020 when the State of California started a stay-at-home order (SAHO), the number and proportion of the U.S. residents under SAHO have increased until April 7 and plateaued afterwards.

The log-linear models used by Joinpoint program and Stata identified similar turning-points. The State in the U.S., total population under SAHO, population-proportion under SAHO and the proportion of positive tests were not linked to daily new cases or deaths. The trend in COVID-19 daily cases reduced after March 23 (*P*<0.001) and further reduced on April 3 (*P*<0.001), which was associated with implementation of SAHO by 10 states on March 23, and the CDC’s recommendation of face-masking, respectively (**Figure 2**). Similarly, there were 2 turning-points in the trends of COVID-19 daily deaths, with a lag time of 10-12 days. Our simulation studies show early-implementation of SAHO would be associated with a significant reduction in daily new cases and deaths while removal of SAHO would be associated with a significant increase in daily new cases and deaths (**Figure 2**). The estimates of Rt based on the 3 reported mean serial-intervals of COVID-19 all started to decline on March 19, when SAHO was first implemented in the U.S., and declined faster after March 23 (**Figure 3**). After a short plateau, Rt continued to decline after April 3 and fell below/around 1.0 on April 13.

**Figure 2.**
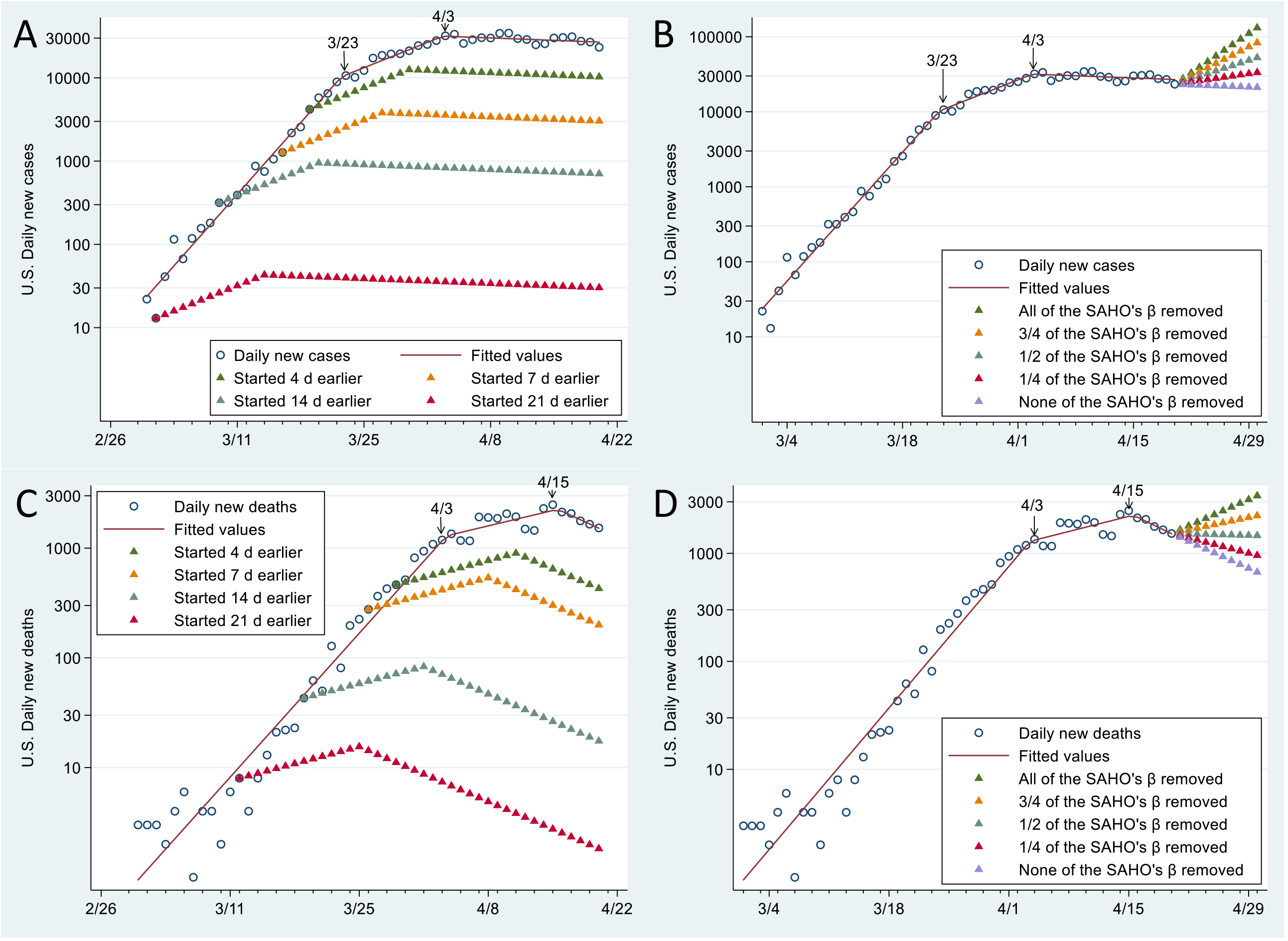
Observed and Simulated Trends in Daily New Cases and Deaths of Laboratory-Confirmed Coronavirus Disease 2019 (COVID-19) in the United States between March 1 and April 30, 2020. The Joinpoint analyses with Poisson variance model show that the 2 turning points of March 23 and April 3 divided the trends in U.S. COVID-19 daily new cases into 3 segments, with the coefficients of 31.69 (95% CI, 26.82 to 36.75, *P*<0.001), 9.75 (95% CI, 7.54 to 12.01, *P*<0.001), −0.90 (95% CI, −1.62 to −0.17, *P=*0.02), respectively. These turning points appeared to link to implementing a stay-at-home order (SAHO) by 10 states on March 23, and the CDC’s face-masking recommendation on April 3. Similarly, the 2 turning points of April 3 and April 15 divided the trends in U.S. COVID-19 daily new deaths into 3 segments, with the coefficients of 25.06 (95% CI, 21.44 to 28.79, *P*<0.001), 5.22 (95% CI, 3.36 to 7.11, *P*<0.001), −7.90 (95% CI, −13.45 to −1.99, *P*=0.01), respectively. The simulated results on early-announcements of SAHO and face-masking recommendation and early-removals of SAHO are shown in A and C, and B and D, respectively. The partial removals of SAHO’s coefficients(ß) may reflex the situations when some of the U.S. states lift the SAHO.

**Figure 3.**
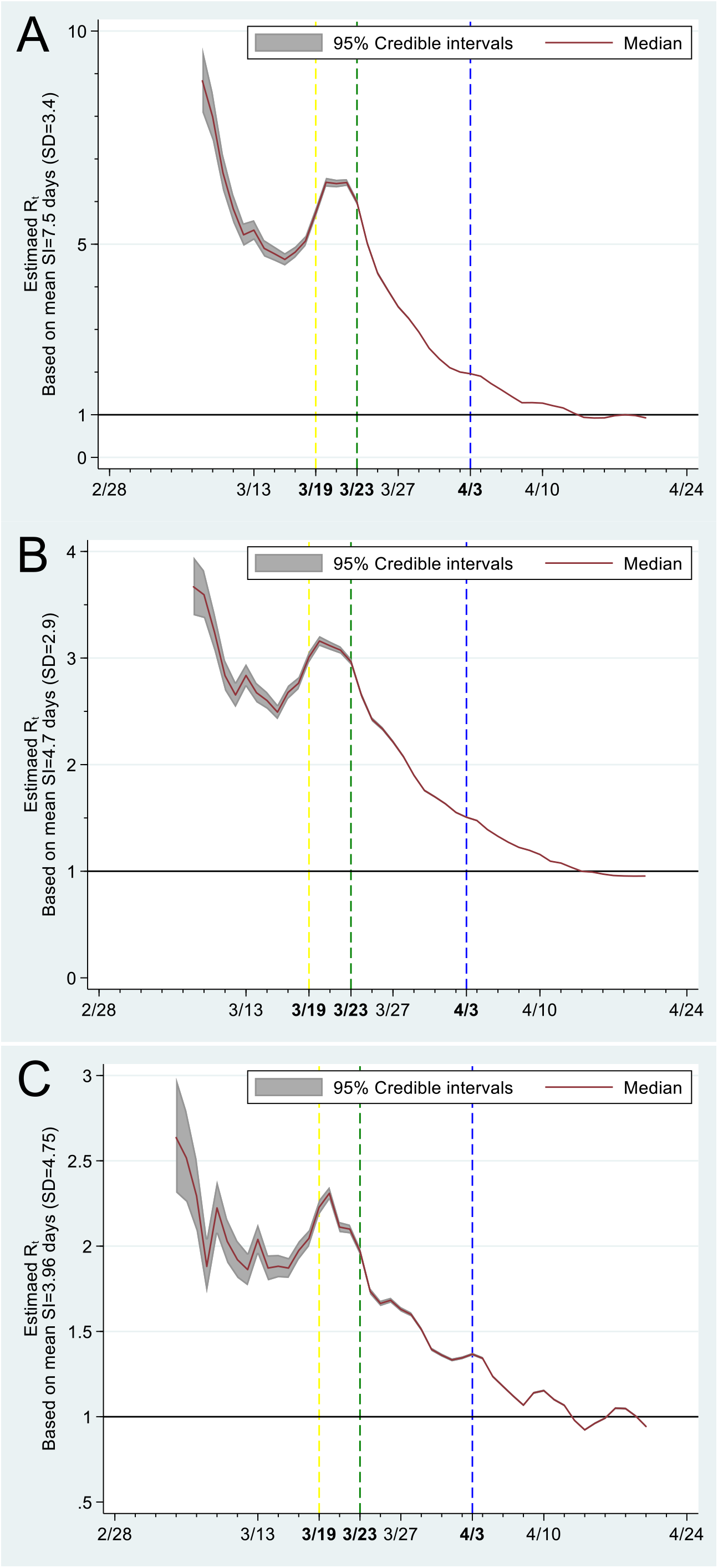
Estimated Effective Reproduction Number (Rt) Based on Laboratory-Confirmed Coronavirus Disease 2019 (COVID-19) Cases in the United States and the Reported Serial Intervals. The effective reproduction number (Rt) was estimated using the previously-reported COVID-19 mean serial intervals (SI) of 7.5, 4.7 and 3.96 days, as well as the corresponding standard deviations (SD). The state-wide stay-at-home-order was first implemented by the state of California on March 19, 2020 (yellow dash line). Ten states had implemented a stay-at-home order by March 23, 2020 (green dash line), affecting 114,047,753 residents (37.45% of the U.S. population). The Centers for Disease Control and Prevention recommended face-masking on April 3, 2020 (blue dash line). These dates were linked to the declines of Rt’s at the times of an increase or plateau of the Rt.

## Discussion

The population under SAHO in the U.S. grew during March 19, 2020 until April 6, 2020, and reached 29,0829,980 (88.6%) by then. The multivariable piece-wise log-linear regression models identified two turning points of COVID-19 daily new cases and the time-varying reproduction number, Rt, in the U.S. They were associated with implementation of SAHO on March 23 and the CDC’s face-masking recommendation on April 3. Similar turning points of COVID-19 daily new deaths were April 3 and April 15, which represented 10 and 12 days of delay, respectively. Simulation on early-implementation and removal of SAHO also reveals considerable impacts on COVID-19 daily new cases and deaths.

### Results in relation to other studies

Several recent works showed that public health interventions including school closure, cordons sanitaire, traffic restriction, social distancing and others, were linked to reduction of Rt and daily incidence of COVID-19.^6-8^ However, these data were mostly from China where the socioeconomic system, and the modalities and extents of public health interventions were significantly different from those in the U.S.^3 4^ Among several modalities of social distancing including school closure, gathering restrictions and restaurant restriction,^4^ the implementing time of SAHO appeared to be the only one matched to the identified turning point (March 23) in this study. Moreover, the Berkeley Interpersonal Contact Study also showed reduction in interpersonal contact in the U.S. between Mach 22 and April 8.^15^ Therefore, our and others’ data together suggest that the implementation of SAHO, which is the strictest social distancing modality, may be required to reach the turning point/timing in trends of new cases. In addition, the studies in China also supports our findings that travel restriction alone would reduce SARS-CoV2 transmission,^6-8^ but additional reduction in transmission in the community (e.g. face-masking in the U.S.) may be required to attenuate or reverse the epidemic trajectory.

The effects of masking on the epidemic of COVID-19 in the U.S. was simulated using the filtering efficacy of masks on influenza.^16 17^ The influence of public intervention on COVID-19 mortalities in the New York State and the U.S. were also simulated using mathematical models and the data of COVID-19, Ebola and influenza viruses.^18^ However, neither of the studies appeared to use laboratory-confirmed cases and COVID-19 based models. Further, neither of the studies simulated the early implementation of SAHO. Therefore, using the population data of laboratory-confirmed cases, we simulated the potential outcomes of implementing SAHO and face-masking recommendation based on the piece-wise (log-linear) models recommended by the guidelines of the U.S. National Center for Health Statistics and other methodological considerations.^19-22^ The simulation studies demonstrate a much smaller scale of COVID-19 in the U.S. when SAHO were implemented earlier, and a concerning reverse of stable downward trends in COVID-19 daily new cases and deaths if being lifted soon. Indeed, the SAHO was not implemented in any of the U.S. states till March 19, when the daily new cases reached 4,190 (crude daily incidence, 12.8 per 1 million) in the U.S.^1^ In contrast, a much more strict SAHO was implemented in China on Jan. 23, when the daily new cases were 259 (crude daily incidence, 0.18 per 1 million) in China and 70 (crude daily incidence, 7.2 per 1 million) in Wuhan.^6 23^ These data suggest an earlier implementation of SAHO in terms of crude daily incidence would be more effective. Consistent with the previous study,^18^ we also show that (early) removal of SAHO would be associated with a second wave or upward trend of COVID-19 daily new cases and deaths. Therefore, caution should be used when considering removal of SAHO.

Rt is one of the most widely used metrics for assessing transmission rate of infectious diseases,^6 18^ and linked to incidence decay with exponential adjustment (IDEA) model and Farr’s law.^24^ However, Rt is difficult to estimated. One of the challenges is the variances in the SARS-CoV2 serial intervals.^11-13^ The reported serial intervals of COVID-19 ranged from 3.96 to 7.5 days, and were first used to rigorously examine the changes of Rt associated with SAHO and face-masking recommendation in the U.S. Two similar turning points were identified in the Rt trends estimated using 3 different serial intervals, and further support the findings discovered using our piece-wise log-linear model.

Geographic differences in COVID-19 incidence and deaths have been reported,^25^ but no quantitative trend analyses were conducted in that report. In light of state difference, we included the States in the U.S. (a variable with 51 categories) as a covariate in our multivariable models, and found it appeared not linked to the trends of daily new cases and deaths. Additional works are needed to better understand the geographic differences in COVID-19 trends.

Many COVID-19 studies were published as preprints. Some of them were of inferior methodological merits and low reporting quality, as discussed before on other fields.^23 26 27^ This observation is concerning, and calls for more collaborative efforts in the efficient review, and timely dissemination of the reports on COVID-19.

### Strengths and weaknesses of this study

The major strength of this study is the use of State-based national data of laboratory-confirmed cases in the analyses.^2^ Moreover, two piece-wise log-linear regression methods were used to rigorously examine trends changes, according to the guidelines on trend analyses of population data and other methodological considerations.^19-22^ Further, the simulation studies provide comprehensive estimates of trends changes linked to early-implementation of SAHO at various time points and early-removal of SAHO with various extents.

Several limitations of this study are noteworthy. First, the positive rates varied among states and by time, suggesting under-testing of the potential patients. The exact COVID-19 case numbers thus are not available, although efforts were made to estimate them using the Johns Hopkins data repository of COVID-19 cases.^28^ Given the data inconsistence we noticed,^1^ such an estimation in our view was not optimal. We were more confident in the reliability of the laboratory-confirmed case numbers. Inclusion of positive-test rate in our models may alleviate the variances in test rate cross the time. Second, there was a lag in COVID-19 reporting,^29^ which may lead to inaccurate estimation of the case numbers. However, the increases in the proportion of the tested population appeared stable in the U.S.^2^ It suggests the lag in reporting may not change significantly as the time changes, and will have minimal impact on the trend analyses. Finally, we did not report the daily incidence. There were no significant changes in the U.S. population during the study-period. The daily new cases of COVID-19 thus should be proportional to its daily incidence in the U.S., but are easier to interpret than daily incidence and were used here.

### Future direction

Future works should be focused on how state level SAHO change the COVID-19 epidemics. This is particularly interesting since state heterogeneity has been reported in 27 European Union states.^30^ As more data, and more-reliable data become available, we would keep looking into the association of SAHO and face-masking policies with the changes in COVID-19 epidemic.

## Conclusions

There were 2 turning points of COVID-19 daily new cases or deaths in the U.S., which appeared to associate with implementation of SAHO and the CDC’s face-masking recommendation. Simulation on early-implementation and removal of SAHO reveals considerable impact on COVID-19 daily new cases and deaths. These findings may inform decision-making of lifting SAHO and face.

## Data Availability

All used data are publicly available.

## Author Contributions

Dr Zhang had full access to all of the data in the study and equally takes responsibility for the integrity of the data and the accuracy of the data analysis.

Concept and design: Xu, Zhang.

Acquisition, analysis, or interpretation of data: All authors.

Drafting of the manuscript: Xu.

Critical revision of the manuscript for important

intellectual content: All authors.

Statistical analysis: Zhang.

Supervision: Zhang.

